# Securitised Health and Zero-Dose Children: Structural and Service-Contact Determinants of Non-Vaccination in Nigeria Paper I

**DOI:** 10.64898/2026.03.02.26347396

**Authors:** Ibrahim Mohammed

## Abstract

**Background:** Zero-dose children, defined as those who have not received a first dose of a diphtheria–pertussis–tetanus (DPT)-containing vaccine, represent one of the sharpest manifestations of inequity in immunisation systems. Nigeria remains one of the largest contributors to the global zero-dose burden, with North-East Nigeria facing intersecting crises of conflict, population displacement, governance fragility, and weakened primary health care. Existing research has largely focused on structural determinants such as poverty, maternal education, and rural residence, with far less attention to relational mechanisms and governance dynamics that shape caregiver decisions.

**Methods:** We conducted a cross-sectional secondary analysis of the 2023 Nigeria Demographic and Health Survey (NDHS) Children’s Recode dataset. Weighted descriptive statistics and survey-adjusted logistic regression models were used to estimate zero-dose prevalence and identify structural and health system–contact determinants among children aged 12–23 months

**Results:** The weighted national zero-dose prevalence was 37.1% (95% CI: 35.2–39.0), meaning more than one in three eligible children had never received a DPT-containing vaccine. The strongest independent predictors of zero-dose status were no ANC visits (aOR = 6.68, 95% CI: 5.52–8.09), no maternal education (aOR = 4.70, 95% CI: 2.89– 7.67), poorest wealth quintile (aOR = 2.79, 95% CI: 1.82–4.27), home delivery (aOR = 1.41, 95% CI: 1.18–1.69), and rural residence (aOR = 1.45, 95% CI: 1.18–1.75). Crude regional disparities were marked but attenuated after adjustment, suggesting that the apparent North-East effect is largely mediated through structural and service-contact pathways.

**Conclusion:** Zero-dose status in Nigeria reflects deep structural exclusion and fragmented early contact with the health system, rather than isolated individual preferences. ANC utilisation and place of delivery emerge as pivotal touchpoints where health systems can either build or erode trust and continuity of care. These findings provide a quantitative foundation for future research exploring relational and contextual mechanisms shaping immunisation exclusion.

## INTRODUCTION

Immunisation remains one of the most effective public health interventions globally, averting an estimated 4–5 million deaths annually (WHO, 2023). Yet despite decades of progress, millions of children continue to miss life-saving vaccines, with persistent gaps concentrated in marginalised, underserved, and fragile settings. Among these children, those classified as “zero-dose” i.e. having received no dose of a DPT-containing vaccine, represent the most extreme manifestation of exclusion from formal health systems (Gavi, 2021; Bosch-Capblanch et al., 2017). Zero-dose prevalence has gained strategic prominence because it signals not only gaps in service delivery, but deeper failures in reach, equity, and system legitimacy.

Globally, zero-dose children are not evenly distributed. They cluster in settings marked by conflict, displacement, poverty, and weak governance (Forshaw et al., 2022). This pattern underscores the limitations of technical framings of immunisation coverage and highlights the need to interrogate structural and relational determinants. Immunisation is not simply an act of uptake; it is shaped by access, socioeconomic resources, historical experience, perception of risk, and trust in institutions (Gilson, 2003; Sheikh et al., 2014). In this sense, zero-dose status is both an epidemiological indicator and a sociopolitical signal.

Nigeria exemplifies this complexity. As one of the largest contributors to the global zero-dose burden, Nigeria embodies stark geographic and socioeconomic disparities. Nationally, zero-dose prevalence remains high despite successive investments in routine immunisation (RI), primary health care (PHC), and campaign-based delivery strategies. Within Nigeria, the North-East stands out as a region of persistent vulnerability, driven by over a decade of insurgency, displacement, infrastructural destruction, and weakened state capacity (Wenham, 2021). Millions have been displaced into camps and dispersed settlements, straining PHC infrastructure and disrupting traditional health-seeking patterns. Health facilities have been destroyed, healthcare workers have fled or been targeted, and movement is restricted by insecurity. In these conditions, immunisation services are often irregular, inaccessible, or distrusted.

Beyond physical barriers, relational dynamics are central to the North-East context. We are theorising that caregivers’ decisions are shaped by perceptions of institutional legitimacy, historical neglect, and contested relationships between communities and state-linked actors. In securitised environments i.e. where health services may be accompanied by militarised presence, surveillance, or coercion, caregivers may interpret immunisation not as benevolent care but as an extension of state power (Elbe, 2006). Such perceptions alter the meanings attached to vaccination, making trust an essential determinant of engagement.

Yet most quantitative research on immunisation in Nigeria focuses on structural determinants such as wealth, maternal education, and residence, without engaging with relational, historical, or governance dynamics (Favin et al., 2012; Rainey et al., 2011). These studies are invaluable for identifying risk patterns but insufficient for understanding why families disengage. Technical interventions such as improved vaccine supply or expanded cold-chain coverage cannot alone address adaptive challenges rooted in trust, identity, and social contract (Heifetz et al., 2009).

This gap creates both a conceptual and operational challenge. Policymakers require more than patterns; they require explanations that can inform relational, community-anchored strategies. Health systems scholars argue that trust, legitimacy, and accountability are foundational to system performance, particularly in fragile settings (Gilson, 2003). However, these constructs are difficult to measure quantitatively, necessitating methodological approaches that integrate statistical patterns with lived experience.

While this study focuses on quantifying structural and health system–contact determinants, understanding relational and governance dynamics remains an important area for future research. Quantitative analysis can illuminate where exclusion is concentrated and which structural and service-contact pathways shape vulnerability. Qualitative inquiry can explain how caregivers interpret these pathways, and why certain groups remain disengaged even when services exist.

Within this framework, zero-dose status functions as a measurable, policy-relevant outcome that anchors the inquiry while pointing beyond itself. Identifying determinants such as lack of ANC contact, home delivery, low maternal education, and rural residence enables the study to map structural exclusion. Yet these determinants also signal relational fracture points which are moments where caregivers either enter the system or turn away from it. ANC, for instance, is not simply a service; it is an encounter that can build or erode trust, respect, and confidence. Home delivery may reflect distance and cost, but also fear, stigma, or negative historical experiences with facilities. Low maternal education can constrain not only knowledge but agency, autonomy, and confidence in institutional actors.

Thus, the adaptive challenge underlying zero-dose vulnerability in North-East Nigeria is multidimensional. It spans material deprivation, geographic exclusion, and relational disruption. It reflects not only the absence of services, but the absence of trust in services. It is shaped by conflict, insecurity, trauma, and fractured governance. This complexity demands a methodological approach capable of capturing multiple layers of causation i.e. structural, relational, and institutional.

Figure 1 presents our conceptual model, illustrating how historical and political factors create a context of securitized health that operates through two key pathways to cause zero-dose status: (1) The Structural Pathway, creating tangible barriers like poverty and poor access; and (2) The Relational Pathway, eroding trust and legitimacy, which also moderates the effect of structural barriers.

**Figure 1.**
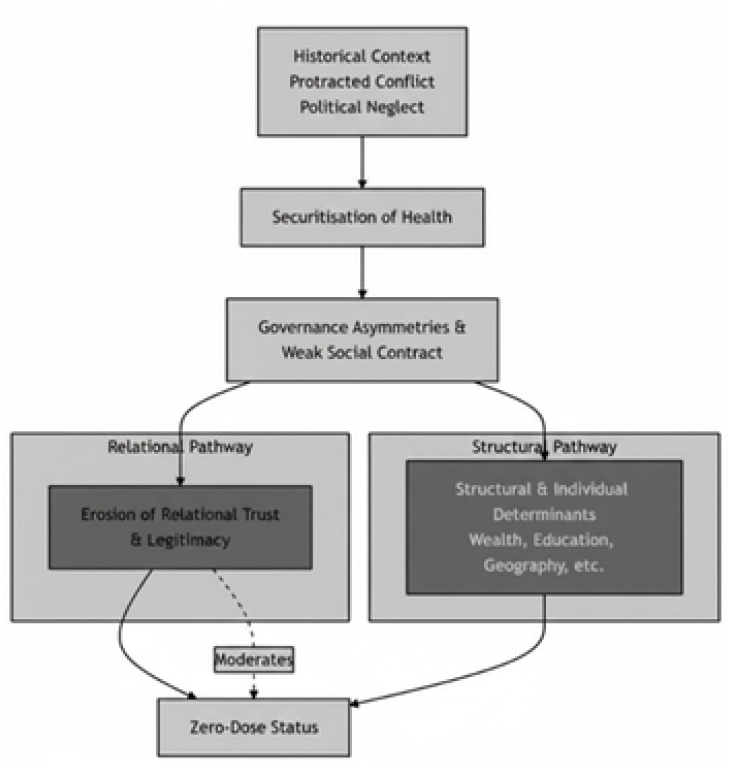
Conceptual model of zero-dose causal factors in a securitised health setting, illustrating how historical and political conditions operate through structural and relational pathways to shape immunisation exclusion in North-East Nigeria.

**Figure 2.**
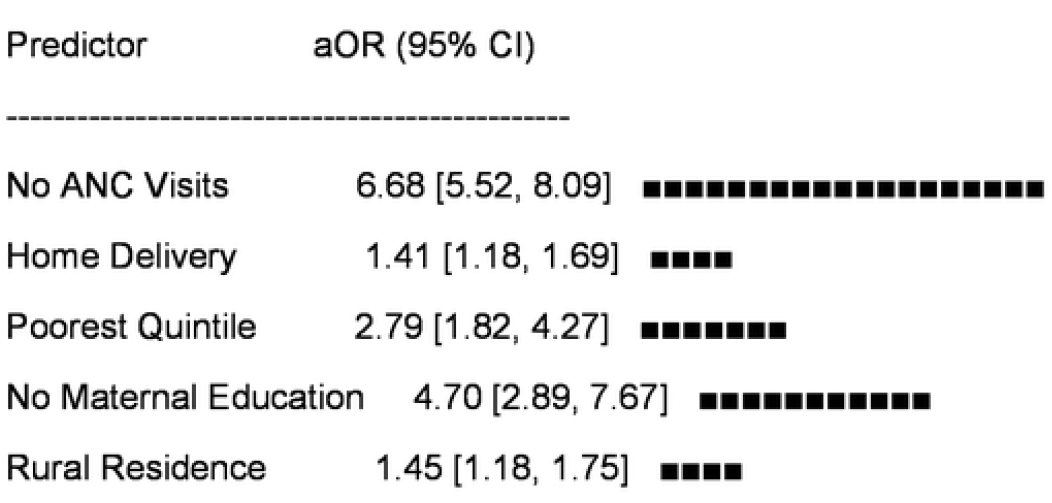
Adjusted odds ratios (aORs) and 95% confidence intervals for predictors of zero-dose status among children aged 12–23 months, Nigeria NDHS 2023. Reference categories are South-East region, richest wealth quintile, maternal higher education, ≥4 ANC visits, facility delivery, and urban residence.

The present study therefore aims to:

1. Quantify structural and health system–contact determinants of zero-dose vaccination among children aged 12–23 months in Nigeria, with explicit attention to North-East patterns; and
2. Generate evidence to inform future qualitative research examining relational and contextual mechanisms underpinning exclusion in North-East Nigeria.
3. By rigorously quantifying structural and service-contact pathways, this study contributes to a clearer epidemiological understanding of immunisation inequity in fragile settings. Ultimately, it seeks to generate insights that support transformative, trust-building interventions for PHC and RI in North-East Nigeria.

## METHODS

### 2.1 Study Design

This study is a cross-sectional secondary analysis of nationally representative survey data from the 2023 Nigeria Demographic and Health Survey (NDHS). The analysis uses the NDHS Children’s Recode (KR) dataset to examine structural and health system–contact determinants of zero-dose vaccination among children aged 12–23 months.

The NDHS employs a stratified two-stage cluster sampling design to produce nationally and regionally representative estimates of demographic, socioeconomic, and health indicators across Nigeria. This design enables population-level inference on immunisation coverage and associated determinants. The present study applies survey weights and design adjustments to account for clustering and stratification, ensuring valid estimation of prevalence and regression parameters.

A cross-sectional analytical approach was selected to quantify the distribution of zero-dose status and assess independent associations with key structural and service-contact variables. The primary outcome is zero-dose status, defined as having received no dose of a DPT-containing vaccine. Explanatory variables include household wealth, maternal education, place of residence, antenatal care utilisation, place of delivery, maternal age, and geographic region. These variables capture dimensions of socioeconomic position, access to services, and early health system contact that have been shown to influence immunisation uptake in low- and middle-income settings.

The analytic strategy focuses on identifying patterns of exclusion and estimating adjusted associations between predictors and zero-dose status using survey-weighted logistic regression models. This approach allows for assessment of whether observed geographic disparities persist after accounting for underlying structural and service-contact factors. By quantifying the relative contribution of these determinants, the study aims to clarify the pathways through which children become disconnected from routine immunisation services within the Nigerian context.

This design is appropriate for addressing the study objectives because it enables rigorous estimation of prevalence and determinants using high-quality, nationally representative data. The findings provide an empirical foundation for understanding structural and health system factors associated with zero-dose status and for informing future research and policy responses aimed at reducing immunisation inequities.

### 2.2.1 Data Source

The quantitative phase draws on the Nigeria Demographic and Health Survey (NDHS) 2023, a nationally representative household survey conducted by the National Population Commission with technical support from ICF International. The Children’s Recode (KR) file contains data on all births to interviewed women in the previous five years, including:

- Child immunisation status
- Maternal characteristics
- Household socioeconomic indicators
- Health service utilisation
- Geographic identifiers

The NDHS uses a two-stage stratified cluster sampling design covering all six geopolitical zones. Survey weights enable valid population inference (Croft, Marshall, & Allen, 2018).

### 2.2.2 Study Population

The analytic sample is restricted to:

- Age: Children aged 12–23 months, ensuring sufficient time for receipt of DPT1.
- Outcome: Non-missing data on all three DPT variables (h3, h5, h7).
- Unit of Analysis: Child, linked to maternal/household characteristics.

This age band reflects cumulative system exposure and aligns with global guidelines for zero-dose measurement (Gavi, 2021).

#### 2.2.3 Key Variables

Primary Outcome

- Zero-dose status (binary): 1 = no recorded or reported DPT1–3; 0 = at least one DPT dose.

Independent Variables

Aligned with the conceptual framework:

- Structural Determinants: Wealth quintile, Maternal education, Residence (urban/rural), Maternal age (continuous)
- Health System Contact Pathways: Antenatal care utilisation (0 / 1–3 / ≥4 visits), Place of delivery (facility/home)
- Contextual Determinant: Region (six geopolitical zones)
- These domains reflect systemic access, sociocultural capability, service touchpoints, and contextual governance (Rainey et al., 2011; Favin et al., 2012).

#### 2.2.4 Statistical Analysis Plan

The analysis proceeds in three stages:

1. **Univariate Analysis:** Weighted prevalence of zero-dose status, Frequency distributions of all predictors
2. **Bivariate Analysis:** Weighted cross-tabulations, Chi-square tests for association, Crude odds ratios
3. **Multivariable Logistic Regression:** Adjusted odds ratios (aORs) with 95% CIs, Survey-weighted estimation using DHS weights

#### Reference categories

South-East, Richest quintile, Higher maternal education, ≥4 ANC visits, Facility delivery, Urban residence

An alpha level of **0.05** is used, consistent with public health standards (Rothman, 2014).

#### 2.2.5 Software

Analysis will be conducted using R (version 4.x) with: haven for data import, dplyr/tidyr for data management, survey for complex design, ggplot2 for visualisation, stats/glm for regression. R is widely used for weighted survey analysis and supports reproducible workflows (Lumley, 2004).

#### 2.2.6 Ethics

The Nigeria Demographic and Health Survey (NDHS) 2023 protocols, including sampling, data collection, and informed consent procedures, were reviewed and approved by the ICF Institutional Review Board and the relevant national ethics bodies in Nigeria. NDHS datasets are fully anonymised prior to release. This secondary analysis involved no contact with human participants and no access to identifiable information, and therefore met criteria for exempt research under standard ethical guidelines. Permission to use the NDHS 2023 Children’s Recode dataset was obtained from the DHS Program.

#### 2.2.7 Sex and Gender Analysis

This analysis focuses on zero-dose status among children aged 12–23 months, using maternal and household characteristics as key explanatory variables. Maternal education, antenatal care utilisation, and place of delivery are understood as gendered processes that reflect women’s autonomy, mobility, and experiences with institutions, rather than neutral individual attributes. While the NDHS 2023 dataset includes child sex, the present analysis did not stratify results by child sex because the primary focus is on maternal and structural pathways. Future research will explore how gender norms, household power relations, and gendered experiences of security shape engagement with antenatal care, childbirth services, and immunisation.

#### 2.2.7 Strobe Reporting Statement

Reporting Guidelines

The quantitative phase of this study follows the Strengthening the Reporting of Observational Studies in Epidemiology (STROBE) guidelines for cross-sectional analyses.

## RESULTS

### 3.1 Sample Characteristics

After restricting the NDHS 2023 KR dataset to children aged **12–23 months** with non-missing DPT vaccination data, the analytic sample comprised **N = 4**,**937** children. Weighted estimates reflect national population parameters. Key characteristics were as follows:

- **Residence:** 63.1% rural; 36.9% urban
- **Wealth Distribution:** Poorest (25.2%), Poorer (20.4%), Middle (18.8%), Richer (19.6%), Richest (16.0%)
- **Maternal Education:** No education (42.0%), Primary (11.3%), Secondary (35.4%), Higher (11.4%)
- **ANC Utilisation:** 0 visits (25.1%), 1–3 visits (15.7%), ≥4 visits (59.2%)
- **Place of Delivery:** Home (54.1%); Facility (45.9%)
- **Regional Distribution:** Largest representation from northern zones

These characteristics reflect a population with substantial structural disadvantage and uneven access to maternal and child health services, aligning with known inequities in Nigeria’s immunisation system.

### 3.2 Weighted Prevalence of Zero-Dose Status

The **weighted national zero-dose prevalence** among children aged 12–23 months was: **37.1% (95% CI: 35.2–39.0)**

This indicates that **more than one in three** eligible children nationally had never received a DPT-containing vaccine. The prevalence exceeds global pre-pandemic averages and underscores persistent system gaps.

### 3.3 Bivariate Analyses

#### 3.3.1 Zero-Dose by Region

| Region | Zero-Dose % |
| --- | --- |
| North Central | 53.9% |
| North East | 35.7% |
| North West | 39.6% |
| South East | 6.9% |
| South South | 12.8% |
| South West | 15.6% |

**Chi-square p <** .**001**

**Interpretation:** Northern regions, especially North Central and North West, carry disproportionate burden. South East performs best, functioning as a benchmark of strong system reach.

#### 3.3.2 Zero-Dose by Wealth Quintile

| Wealth Quintile | Zero-Dose % |
| --- | --- |
| Poorest | 56.8% |
| Poorer | 51.9% |
| Middle | 39.1% |
| Richer | 22.5% |
| Richest | 6.6% |

**Chi-square p <** .**001**

**Interpretation:** A steep socioeconomic gradient suggests material deprivation as a central determinant of immunisation exclusion.

#### 3.3.3 Zero-Dose by Maternal Education

| Education Level | Zero-Dose % |
| --- | --- |
| No Education | 59.1% |
| Primary | 36.8% |
| Secondary | 18.5% |
| Higher | 5.3% |

**Interpretation:** Maternal education is one of the strongest protective factors, with a tenfold difference between highest and lowest categories.

#### 3.3.4 Zero-Dose by Residence

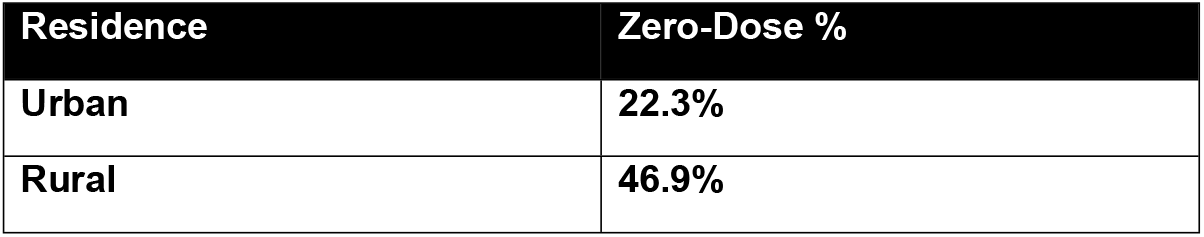

**Chi-square p <** .**001**

**Interpretation:** Rural residence more than doubles the risk of exclusion, indicating spatial access barriers.

#### 3.3.5. Zero-Dose by ANC Utilisation

3.3.5 Zero-Dose by ANC Utilisation
| ANC Visits | Zero-Dose % |
| --- | --- |
| 0 Visits | 77.1% |
| 1-3 Visits | 37.2% |
| $\geq 4$ Visits | 19.6% |

**Chi-square p <** .**001**

**Interpretation:** ANC contact represents a critical system gateway. Absence of ANC corresponds with the highest observed exclusion level in the entire dataset.

#### 3.3.6 Zero-Dose by Place of Delivery

| Place of Delivery | Zero-Dose % |
| --- | --- |
| Facility | 16.3% |
| Home | 53.9% |

**Chi-square p <** .**001**

**Interpretation:** Home delivery triples the prevalence of zero-dose status, underscoring childbirth as a decisive system interface.

### 3.4 Multivariable Logistic Regression

A survey-weighted logistic regression model examined independent predictors of zero-dose status. Reference categories were selected to represent optimal access:

- **Region:** South East
- **Wealth:** Richest
- **Education:** Higher
- **ANC:** ≥4 visits
- **Delivery:** Facility
- **Residence:** Urban

#### Key Adjusted Odds Ratios (aORs)

| Predictor | aOR 95% CI | p-value |
| --- | --- | --- |
| Poorest wealth quintile | 2.79 [1.82, 4.27] | <.001 |
| Maternal education (none) | 4.70 [2.89, 7.67] | <.001 |
| ANC (0 visits) | 6.68 [5.52, 8.09] | <.001 |
| Home delivery | 1.41 [1.18, 1.69] | <.001 |
| Rural residence | 1.45 [1.18, 1.75] | <.001 |
| North Central region | 1.58 [1.13, 2.21] | .007 |
| North East region | 0.81 [0.57, 1.15] | 0.24 |

### 3.5 Forest Plot Visualization

The adjusted odds ratios from the multivariable model are visualized in Figure 3, providing immediate interpretation of effect sizes and precision across all predictors.

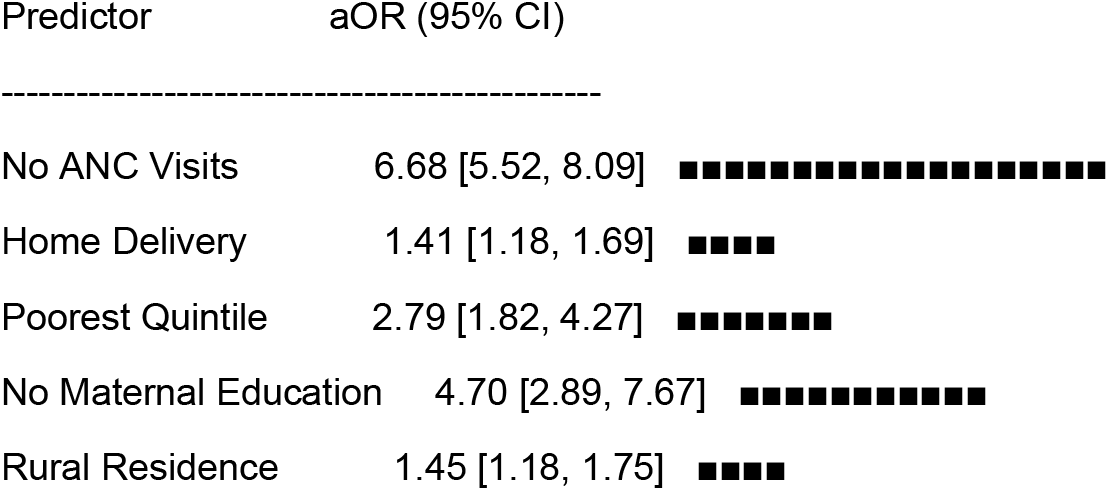

### 3.5 Interpretation of Multivariate Findings

#### 3.5.1 Structural Inequity

Wealth and maternal education remain powerful, independent predictors. The four-fold higher odds among children of mothers with no education suggests deep sociocultural and informational barriers, reinforcing education as a proxy for empowerment and system navigation.

#### 3.5.2 System Contact Pathways

ANC emerges as the single strongest determinant. The near seven-fold increase in odds for children whose mothers had zero ANC visits suggests early system disengagement cascades into immunisation exclusion. Home delivery further amplifies this pathway.

#### 3.5.3 Geography and Context

While the North East showed high crude prevalence, its effect **disappeared after adjustment**, indicating that structural deprivation and low system contact, and not region itself, drive exclusion. These finding shifts focus away from geography per se toward underlying mechanisms.

#### 3.5.4 Protective Factors

Maternal age exhibited a small protective effect, suggesting that younger mothers may face greater barriers related to autonomy, mobility, or social support.

## DISCUSSION

This study provides robust empirical evidence that zero-dose vaccination in Nigeria is not a random occurrence but a patterned manifestation of structural inequity and fragmented maternal–child health system contact. Using a nationally representative dataset, the analysis demonstrates that exclusion from immunisation is deeply embedded in socioeconomic disadvantage, educational deprivation, and missed early engagement with formal health services. These findings reinforce a growing body of literature showing that immunisation inequalities mirror broader social hierarchies and structural vulnerabilities rather than individual preferences or isolated service failures (Bosch-Capblanch et al., 2017; Favin et al., 2012; Rainey et al., 2011).

A central contribution of this analysis is the clear identification of **ANC utilisation** as a pivotal determinant of zero-dose status. The near seven-fold increase in odds among children whose mothers received no ANC suggests that ANC is not simply a clinical encounter, but a gateway to system awareness, continuity of care, and trust-building. This aligns with evidence that ANC predicts both service familiarity and subsequent uptake of preventive interventions (Mutua et al., 2020). Home delivery, which was similarly associated with markedly higher zero-dose prevalence, reinforces the idea that childbirth functions as a decisive touchpoint at which health systems can either enable or foreclose future engagement. These findings underscore the importance of strengthening continuity between maternal health and immunisation platforms within a primary healthcare (PHC) framing, consistent with integrated models advocated by WHO (2023).

The strong independent effects of **wealth and maternal education** highlight the deep socioeconomic stratification of immunisation access. The steep gradient observed from fewer than 7% zero-dose prevalence in the richest quintile to nearly 57% in the poorest, suggests that financial deprivation, opportunity costs, and constrained mobility materially limit health-seeking behaviours. Maternal education, functioning as a proxy for health literacy, empowerment, and decision-making capacity, amplifies these inequalities. These findings echo prior analyses across LMICs demonstrating that socioeconomic inequities remain the dominant driver of immunisation coverage gaps (Wiysonge et al., 2012; Hosseinpoor et al., 2016). Interventions must therefore address not only service supply, but also the enabling conditions that allow caregivers to access and trust those services.

A particularly notable insight concerns the role of **geographic region**. Although the North East appeared highly burdened in crude analyses, consistent with its conflict-affected and fragile system profile, the effect dissipated in the multivariable model. This suggests that regional differences are largely mediated by structural deprivation, reduced ANC and maternity service utilisation, and rural residence, rather than geography per se. While the quantitative results deprioritise region as an independent determinant, they simultaneously elevate the need to understand why structural and relational disadvantages cluster so strongly in this region. Conflict, displacement, fragmented governance, and eroded institutional legitimacy, these are factors not captured in DHS variables but likely shape caregivers’ perceptions of safety, trust, and acceptability of formal services. These are precisely the kinds of mechanisms that qualitative inquiry is designed to surface (Gilson, 2003; Sheikh et al., 2014).

Quantitative results identify where exclusion is concentrated and which pathways matter most and should be followed up with a qualitative phase which will address how lived experience, relational trust, and institutional histories shape disengagement. For instance, qualitative interviews may reveal whether avoidance of ANC or facility delivery reflects fears of mistreatment, gendered power constraints, security risks, or perceptions of state institutions as instruments of surveillance rather than care, a dynamic noted in securitised health contexts (Wenham, 2021). These relational mechanisms cannot be inferred from statistical patterns alone but are essential to designing interventions that resonate with community realities.

Taken together, the findings point toward a paradigm shift: zero-dose exclusion in Nigeria is less a function of vaccine hesitancy or logistical gaps and more a symptom of deeper systemic fractures across social, economic, and relational domains. Addressing this challenge requires moving beyond technical solutions and toward adaptive strategies that strengthen trust, reduce structural barriers, and rebuild continuity between maternal health and immunisation services. By providing robust empirical modelling of structural and service-contact determinants, this study offers a quantitative foundation for future work examining relational and governance dimensions of immunisation exclusion.

## LIMITATIONS

This study has several important limitations that should guide interpretation and the next phase of inquiry. First, the quantitative analysis relies on **proxy indicators** rather than direct measures of the underlying constructs. Variables such as maternal education and ANC utilisation approximate sociocultural capability and system engagement but do not capture autonomy, decision-making power, perceived quality of care, or trust, these are factors that are likely central to immunisation behaviour. Second, the NDHS immunisation indicators partially rely on **maternal recall**, introducing potential recall and misclassification bias, particularly among caregivers with low literacy or limited exposure to health documentation. Third, the dataset **lacks variables on conflict exposure, displacement, insecurity, or governance**, which may confound the relationship between geographic context and zero-dose status, especially in the North-East. Fourth, the cross-sectional design precludes causal inference; associations identify patterns but cannot determine temporal sequencing or directionality. Finally, children who have died or migrated are not represented in the dataset, potentially underestimating exclusion in the most fragile settings.

These limitations highlight the need for future research exploring relational and contextual mechanisms not directly captured in DHS datasets.

## CONCLUSIONS

This study provides compelling evidence that zero-dose exclusion in Nigeria reflects deep structural inequities and fragmented early contact with the health system. Wealth, maternal education, ANC utilisation, home delivery, and rural residence emerged as powerful determinants, while the apparent regional effect diminished once these structural and contact pathways were accounted for. These findings suggest that immunisation outcomes are shaped less by geography itself and more by clustered deprivation and relational distance from formal health systems. The results underscore the need for interventions that strengthen maternal–child health continuity, reduce socioeconomic barriers, and rebuild trusted points of engagement early in the care pathway.

This nationally representative quantitative analysis identifies key structural and service-contact determinants shaping zero-dose exclusion in Nigeria and provides an empirical foundation for future research examining relational and governance dimensions of immunisation inequity. Quantitative patterns identify where vulnerability is concentrated, while qualitative inquiry will illuminate why caregivers disengage and how trust, legitimacy, and lived experience shape immunisation decisions. By linking structural determinants to relational dynamics, this study lays the foundation for transformative, equity-oriented strategies capable of reducing zero-dose exclusion in fragile settings and advancing Nigeria’s progress toward universal immunisation coverage.

## Data Availability

The data underlying the results presented in this study are publicly available from the Demographic and Health Surveys (DHS) Program website at https://dhsprogram.com/data/available-datasets.cfm. The specific dataset used is the 2023 Nigeria Demographic and Health Survey (NDHS) Children’s Recode (KR) file, which contains information on immunisation status and maternal and household characteristics for children born in the five years preceding the survey. Access to the NDHS 2023 dataset requires registration with the DHS Program and agreement to their standard terms of use, including restrictions on redistribution and requirements to cite the source appropriately. Researchers who meet the DHS Program’s eligibility criteria can request access to the anonymised dataset for secondary analysis. No custom or proprietary data were generated or analysed in this study.

## AUTHOR CONTRIBUTIONS (CReDiT)

Ibrahim Mohammed: Conceptualization; Methodology; Data curation; Formal analysis; Visualization; Writing – original draft; Writing – review and editing; Project administration.

## DATA AVAILABILITY STATEMENT

This study is based on secondary analysis of the 2023 Nigeria Demographic and Health Survey (NDHS) Children’s Recode dataset. These anonymised data are available from the DHS Program (https://dhsprogram.com) upon reasonable request and registration.

## ICMJE AUTHORSHIP STATEMENT

All authors attest they meet the ICMJE criteria for authorship.

## DECLARATION OF GENERATIVE AI USE

During the preparation of this work, the author used generative AI tools to support language refinement, organisation of content, and identification of gaps in alignment with journal author guidelines. After using these tools, the author reviewed, edited, and verified all content and takes full responsibility for the accuracy and integrity of the manuscript.

